# MRGPRX2-expressing mast cells are increased in the GI tract of individuals with active inflammatory bowel disease and hereditary α-tryptasemia

**DOI:** 10.64898/2025.12.23.25342823

**Authors:** Michelle Galeas-Pena, Jonathan J. Lyons, Dhana Llivichuzhca-Loja, Steve Everman, Ghina Yaghi, Katelyn White, Sangita Sutradhar, Tanya O. Robinson, Anna H. Owings, Neha S. Dhaliwal, Alexander Carlye, Liza Konnikova, Hydar Ali, Sarah C. Glover

**Affiliations:** Department of Medicine, Section of Gastroenterology. Tulane University School of Medicine. New Orleans, LA; Division of Allergy & Immunology, Department of Medicine, University of California San Diego, La Jolla, CA; Veterans Affairs San Diego Healthcare System, La Jolla, CA; Departments of Pediatrics, Immunobiology and Obstetrics, gynecology and reproductive sciences, Yale University School of Medicine, New Haven, CT; Division of Digestive Diseases, Department of Medicine, University of Mississippi Medical Center, Jackson, MS; Department of Basic and Translational Sciences, University of Pennsylvania, School of Dental Medicine, Philadelphia, PA

**Author notes:** Correspondence: Sarah C. Glover, DO Section of Gastroenterology and Hepatology Department of Medicine Tulane University School of Medicine 131 S Robertson St. Mail Code 8035 New Orleans, LA 70119. Hydar Ali, Ph.D. Department of Basic and Translational Sciences, University of Pennsylvania, School of Dental Medicine 240 South 40th Street Philadelphia, PA 19104.

**Keywords:** Mast cells, MRGPRX2, inflammatory bowel disease, hereditary alpha-tryptasemia, gastrointestinal tract, mass cytometry, SIGLEC8

## Abstract

Hereditary α-tryptasemia (HαT), defined by increased *TPSAB1* copy number and elevated basal serum tryptase, is associated with mast cell (MC)–mediated symptoms, yet its role in gastrointestinal disease remains unclear. Because intestinal MCs express the non-IgE–dependent activation receptor MRGPRX2, we investigated whether MRGPRX2 expression is altered in individuals with HαT and inflammatory bowel disease (IBD). We genotyped 854 biobanked IBD samples, performed spatial transcriptomics on descending colon tissue (n = 4 HαT; n = 4 non-HαT), and analyzed small-intestinal biopsies by mass cytometry (CyTOF; n = 5 HαT; n = 9 controls). Across these complementary platforms, HαT was associated with increased gastrointestinal MC abundance and higher expression of activation markers including CD203c, LAMP-1, and SIGLEC8. Both spatial transcriptomics and ddPCR demonstrated significantly increased MRGPRX2 and SIGLEC8 transcripts in IBD samples from individuals with HαT compared with matched non-HαT IBD cases. These findings suggest that enhanced MRGPRX2 expression and associated MC activation may contribute to gastrointestinal symptoms in HαT, particularly in the context of IBD. As interest in precision immunogenetics grows, defining MC phenotypes linked to α-tryptase copy number may help refine diagnostic evaluation and identify patients who could benefit from emerging mast cell–targeted therapeutic strategies in the context of IBD.

## 1 Introduction

Hereditary α-tryptasemia (HαT) is classified as an autosomal dominant genetic trait prevalent in western populations (1,2). It is characterized by elevated basal serum tryptase (BST) of >8 ng/ml resulting from increased copy number of the *TPSAB1* gene encoding α-tryptase (3–7). Severe symptoms can be associated with this trait and include anaphylaxis, gastrointestinal symptoms, and food and chemical sensitivity (8,9).

Hereditary α-tryptasemia has also been shown to influence mast cell–associated biology in certain contexts, although its clinical expression is heterogeneous (7,10,11). Several studies have reported an overrepresentation of HαT among individuals with clonal mast cell disorders such as systemic mastocytosis and have described a gene-dose effect on BST together with increases in mediator-related symptoms (12–15). Tissue-based analyses have further shown increased duodenal mast cell numbers in individuals with HαT relative to controls (15,16) (10,14), suggesting that α-tryptase gene dosage may affect mast cell homeostasis in the gastrointestinal tract. These observations, raise the possibility that HαT could modulate mast cell–dependent pathways in specific biological or clinical settings.

In humans, MCs are the primary source of tryptases. There are several isoforms of tryptase, such as alpha (α-), beta (β-), gamma (γ-), and delta (δ-), with secreted isoforms being α- and β-tryptases, which are the only ones associated to clear physiological and pathological outcomes, hence the focus of this study(2,11).

Mature tryptases assemble into tetramers that can be either heterotetrameters (α:β) or α-tryptase (α:α) or β-tryptase (β:β) homotetramers (10,12,15). While α-tryptase homotetramers have negligible proteolytic activity, α:β-heterotetramers, but not β-tryptase homotetramers, render MCs susceptible to vibration-triggered degranulation. This occurs through cleavage of the α-subunit of the mechanosensing adhesion G protein coupled receptor (GPCR) epidermal growth factor (EGF)-like module-containing mucin-like hormone receptor-like 2 (EMR2) encoded by *Adhesion GPCR E2* (*ADGRE2*) (2). This may help explain why some individuals with HαT display cutaneous symptoms such as flushing and erythema in response to vibratory stimuli (17).

Mature tryptases have been shown to contribute to immediate hypersensitivity in a humanized mouse model of anaphylaxis; neutralization of their enzymatic activity was shown to limit IgE-mediated temperature drop in this context (18). Moreover, MC activation and degranulation can be heterogeneous, depending on specific tissue localization and IgE production(19). Some MCs of mucosal origin express tryptase only, and connective-tissue MC canonically express tryptase, chymase, and carboxypeptidase A3 (CPA3)/cathepsin G, with their relative abundance differing by site (20). Single-cell atlases show that MCs adopt tissue-specific transcriptional programs and cluster primarily by organ; in mice, conserved mucosal-type vs MrgprB2⁺ connective-tissue–type signatures are evident, with distinct programs observed in human tissues (20,21). However, in the lung, scRNA-seq of sorted human MCs finds a predominance of tryptase-expressing MCs with variable *CMA1*/*CTSG* and high *CPA3* mRNA. These data exemplify that heterogeneity is likely a continuum rather than a representation of discrete subtypes, and also suggests that there may be plasticity within this compartment that may be modified by host or environmental factors (22).

Because β-tryptase-induced MC degranulation is sensitive to pertussis toxin, it is postulated that a Gαi-protein coupled, like Mas-Related GPCR--X2 (MRGPRX2), is involved in this autocrine mechanism (11,23). MRGPRX2 is a GPCR that is predominantly expressed in connective tissue MCs (24) and, while largely studied in the skin, it is also expressed in intestinal MCs (8,25). This receptor is involved non-IgE-mediated immediate hypersensitivity reactions (26). Cationic ligands including host defense antimicrobial peptides (HBDs), neuropeptides, wasp venom, neuromuscular-blocking drugs, and fluoroquinolone antibiotics have been shown to activate MCs via MRGPRX2 *in vitro* (24,27,28). Of particular clinical relevance is the association between HαT and Hymenoptera venom-mediated anaphylaxis (HVA) where patients with HαT present with more severe HVA than those without (3,10,15,29,30).

Recent studies have reported that certain MRGPRX2 agonists are upregulated in colonic tissues of inflamed ulcerative colitis (UC) patients compared to those with inactive disease (25). MRGPRX2 has also been observed to increase MC-mediated release of histamines which may contribute to gastrointestinal diseases such as irritable bowel syndrome (IBS) and intestinal pain (31) (32). Additionally, we have observed similarities in clinical manifestations and increased intestinal epithelium pyroptosis in patients with HαT and those with quiescent IBD (8). A recent publication has also associated HαT with refractory Celiac disease in individuals otherwise histologically responding to a gluten-free diet (33). Therefore, the possibility of an increased presence or expression of MRGPRX2 in the gastrointestinal (GI) tract of symptomatic individuals with HαT requires examination.

Herein, we show that MRGPRX2 is increased in GI mucosal biopsies from individuals with inflammatory bowel disease (IBD) and HαT. Furthermore, we demonstrate that MRGPRX2+ small intestinal (SI) MCs have a higher expression of CD63, CD203c, and LAMP1 compared to MCs that do not express MRGPRX2.

## 2 Materials and Methods

### Study participants and sample availability

DNA samples to determine α-tryptase copy number variant were kindly provided by the IBD biobanks from University of Pennsylvania and Mayo Clinic. Using the same participant pool from the University of Pennsylvania IBD biobank, we obtained tissue matching location (large intestine) and severity (moderate) for transcriptomics analysis was obtained from the IBD biobank from the University of Pennsylvania. Additional tissue samples to perform cytometry by time of flight (CyTOF) were obtained from participants who were given informed consent from our approved IRB research protocols at NIAID (NCT01164241, NCT00852943, UMMC (IRB 2019-0082).

Samples from our previous study (8) were also used as disease controls: patients with quiescent Crohn’s disease (CD) – defined as any subject with a previous CD diagnosis, who had no evidence of active inflammation on ileal biopsy at the time of the study – who did not have HαT, provided informed consent and were enrolled on the same IRB-approved protocols. We collected Clinical and histopathological information from the respective electronic medical record systems at University of Mississippi Medical Center (UMMC) in accordance with approved human participant protocols. Additional RNA samples kindly provided by Dr. Therrien at Beth Israel Deaconess from patients with and without HαT (n=16 and 19, respectively) without IBD was used for MRGPRX2 expression validation.

A detailed summary of participant numbers, disease phenotypes, tissue sources, and analytic platforms for each cohort is provided in Table 1 and illustrated in Figure 1A. Data is publicly available at https://github.com/nuzla/-GEO_HaT_Xenium_Submission

**Figure 1.**
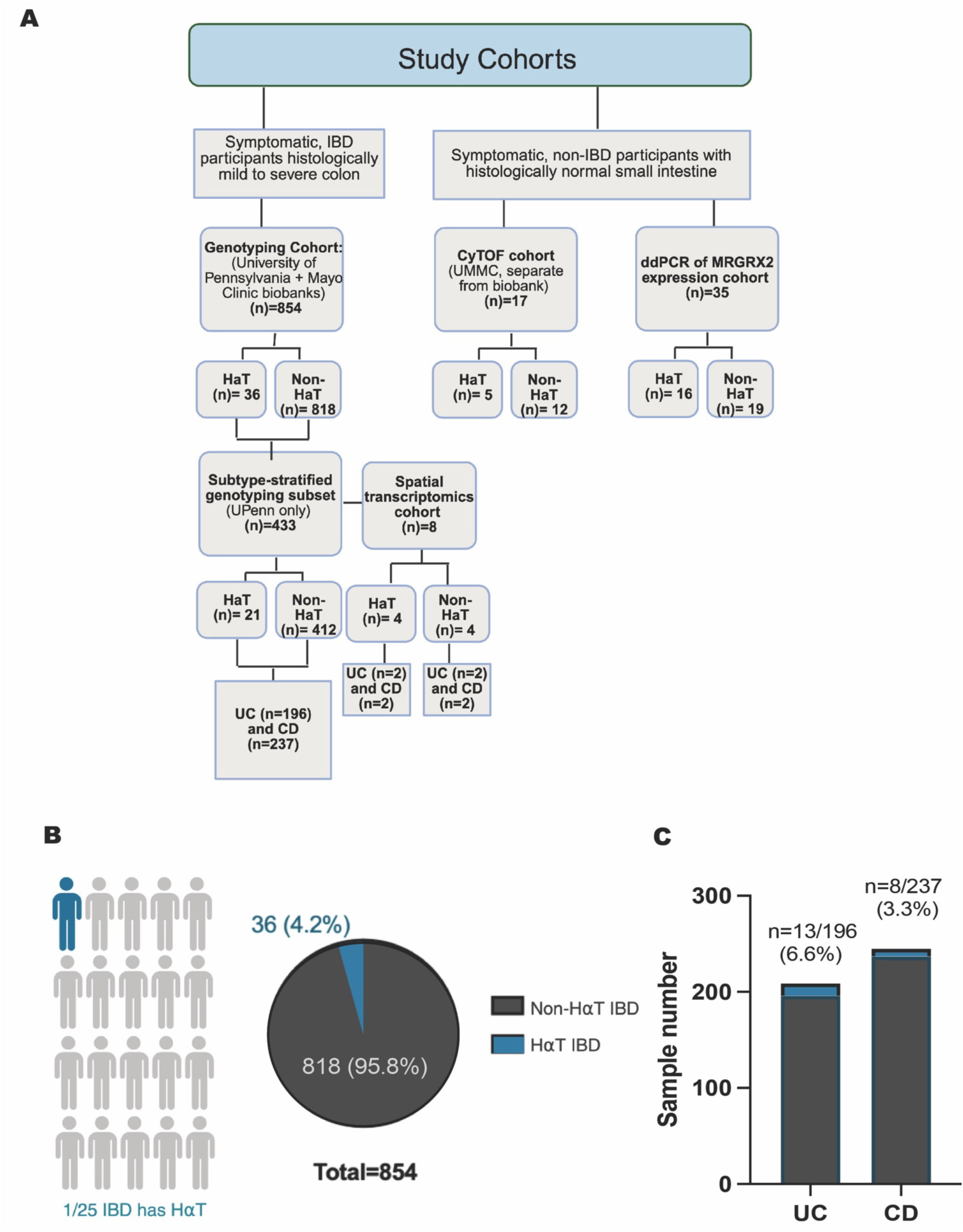
Prevalence of HαT in biobanked IBD samples and distribution across UC and CD. A) Study cohort description. B)TPSAB1 copy-number variation was assessed by digital droplet PCR (ddPCR) in 854 DNA samples from the University of Pennsylvania and Mayo Clinic IBD biobanks. Overall, 4.2% (36/854) of samples were positive for hereditary α-tryptasemia (HαT). C) Among the subset with known IBD subtype (n = 433), HαT prevalence was 13/196 in ulcerative colitis (UC) and 8/237 in Crohn’s disease (CD). Proportions were compared using a two-sided Fisher’s exact test; effect sizes are reported as risk difference with 95% CI. Panel A created with BioRender under institutional license.

**Table 1.** Summary of the study cohorts included in genotyping, spatial transcriptomics, and CyTOF analyses. The large ddPCR genotyping cohort (n=854) identified individuals with hereditary α-tryptasemia (HαT) and provided the basis for disease-stratified analyses and selection of matched samples for downstream molecular studies. Spatial transcriptomics was performed on a balanced subset of eight severe IBD samples (4 HαT, 4 non-HαT) from the descending colon. CyTOF was performed on an independent cohort of symptomatic but histologically normal small intestinal biopsies (n=17). This table clarifies the relationship between the genotyped cohort and the molecular datasets.

| Cohort / Analysis | N Samples | HaT (n) | Non-HaT (n) | Clinical Phenotype | Tissue Type | Analytic Platform | Notes |
| --- | --- | --- | --- | --- | --- | --- | --- |
| **Genotyping Cohort** (University of Pennsylvania + Mayo Clinic biobanks) | 854 | 36 | 818 | IBD (UC, CD) and non-phenotyped biobank submissions | DNA from biobanked whole blood | ddPCR (TPSAB1 CNV) | Used to determine HaT prevalence and to identify matched samples for downstream molecular analysis. |
| **Subtype-stratified genotyping subset** (UPenn only) | 433 | 21 | 412 | UC (n=196) and CD (n=237) | DNA | ddPCR (TPSAB1 CNV) | Provided disease-specific HaT frequency estimates (UC: 13/196; CD: 8/237). |
| **Spatial transcriptomics cohort** | 8 | 4 | 4 | Severe IBD: UC (n=4), CD (n=4) — balanced across HaT and non-HaT | Descending colon | 10x Visium Spatial Transcriptomics + scRNA-binning | Selected from genotyped cohort; used to evaluate MC abundance and MRGPRX2 expression patterns. |
| **CyTOF cohort** (UMMC, separate from biobank) | 17 | 5 | 12 | Symptomatic, non-IBD participants with histologically normal small intestine | Small intestine | Mass cytometry (CyTOF) | Independent functional cohort used to assess MC phenotypes associated with HaT. All samples H&E-normal. |
| **ddPCR of MRGRX2 expression cohort** | 35 | 16 | 19 | Symptomatic, non-IBD participants with histologically normal small intestine | Small intestine | ddPCR | Used to evaluate MRGPRX2 expression independent of spatial transcriptomics |

### Tryptase genotyping

Droplet digital PCR was performed as described (1) to determine tryptase genotypes of all study participants on a research basis in the laboratory of Dr. Lyons at University of California, San Diego (UCSD) or using a clinical laboratory (Gene by Gene, Houston, TX).

### Total basal serum tryptase quantification

Total basal serum tryptases (BST) levels were measured in a subset of participants using a commercially available ImmunoCAP assay (Thermo Fisher Scientific, Phadia AB, Uppsala, Sweden) and performed in a CLIA-certified laboratory (Mayo Clinic, Rochester, NY). BST values were not required for cohort inclusion, as they are typically obtained only when clinically indicated and are not routinely measured in asymptomatic controls or in biobanked specimens collected for unrelated clinical purposes. Classification of hereditary α-tryptasemia (HαT) for the present study was therefore based exclusively on *TPSAB1* copy number determined by digital droplet PCR (ddPCR), the definitive diagnostic assay for HαT. BST levels were not used as an analytical variable in the molecular studies.

### Isolation of lamina propria mononuclear cells and CyTOF

Single cell suspensions were made from small intestine biopsies (n = 17; 5 HαT and 9 controls) from symptomatic, non-IBD participants with histologically normal mucosa were analyzed by mass cytometry. Samples were collected in 90% FBS with 10% DMSO and slow frozen as described (34). Thawed small intestinal tissue samples were digested overnight on a shaker at 37° C in complete RPMI media with 2 µL of collagenase and 2 µL of DNase per 12.5 mL media. Undigested material was filtered out using a 100 µM filter. Single cells were resuspended in CyTOF staining buffer and counted. Cells, 1 x 10^6^/sample were prepared for CyTOF according to the Fluidigm protocol (8,34) with minor modifications. Cells were stained with Rh103 for viability, washed, blocked with Fc-Block and incubated with the cocktail of metal-coupled surface antibody for 30 min. Cells were then resuspended in FOXP3 fixation and permeabilization buffer (Invitrogen) for 45 min and stained with intracellular antibody cocktail for 45 min. They were finally fixed in 1.6% formaldehyde and stained with Ir-DNA intercalator solution. Cells were resuspended in CAS solution 1:10 dilution of EQ beads and run on a Helios CyTOF machine (Fluidigm) at the HMS CyTOF Core at Yale. All antibodies were obtained from either Fluidigm or from the Harvard CyTOF antibody core. Data were analyzed using premium CyTOBANK cloud-based software. Statistical analysis was performed using GraphPad Prism 7 software (GraphPad Software, San Diego, CA). All samples were processed, stained, barcoded, and acquired in a single CyTOF run, eliminating acquisition-related batch effects and no additional computational batch correction was required. Data preprocessing included bead normalization, debarcoding, and gating in FlowJo and Cytobank. Two mast cell populations (CD45⁺ Lin⁻ c-KIT⁺ FcεRI⁺ Tryptase⁺) were identified. Marker intensities were arcsinh-transformed (cofactor = 5). Group differences (HαT vs non-HαT; MRGPRX2⁺ vs MRGPRX2⁻) were evaluated using Wilcoxon rank-sum test. For all comparisons, effect sizes (Cohen’s d) and 95% confidence intervals were calculated. Because only a small, predefined set of activation markers was examined, Benjamini–Hochberg FDR correction was applied when >3 markers were tested simultaneously. See Supplemental Table 2 for antibody and metal list and Supplemental Figure 3 for gating strategy.

### Digital Droplet PCR

To determine expression of *MRGPRX2*, we used a premade probe (BioRad Cat# 10031252) and followed manufacturer’s instructions. The assay was run in a BioRad automated droplet generator and BioRad QX600 droplet digital PCR system with BioRAD ddPCR reagents at the manufacturer recommended concentrations. Results were analyzed with BioRad Quantasoft Software (version 2.1).

### Spatial Transcriptomics

Frozen descending colon biopsies from 4 individuals with HαT and 4 matched non-HαT IBD controls (tissue type and severity) from the University of Pennsylvania IBD biobank were analyzed using the 10x Genomics Xenium platform as described (35). Xenium Explorer v3.1 was used for imaging, transcript alignment, and feature selection. Raw count matrices were imported into R (v2023.06.1+524) and processed with Seurat v5.3.0. Low-quality cells were removed (<200 genes or >20% mitochondrial RNA). Normalization was performed using SCTransform with mitochondrial regression, followed by PCA on the top 3,000 HVGs. Clustering was performed using the Louvain algorithm (resolution = 0.5). Marker genes were identified using a Wilcoxon rank-sum test with Benjamini–Hochberg FDR correction (adjusted p < 0.05 and log₂FC > 0.25). Pseudobulk counts were generated by summing transcripts per cluster × sample, and differential expression between HαT and non-HαT conditions was evaluated using DESeq2 (v1.42.1) with BH-adjusted FDR < 0.05. Effect sizes (log₂FC and Cohen’s d with 95% CIs) were computed at the pseudobulk level. All visualizations were created using ggplot2 (v3.5.2), cowplot (v1.1.3), and Seurat plotting functions. All packages used are listed in Supplemental Table 3.

### Seurat and pseudobulk analysis

Analyses were performed in R (v4.3) with Seurat v5. Low-quality cells were removed using distribution-aware filters (Tukey fences for counts, features, and mitochondrial percentage). HVGs (up to 3,000 genes/assay) were identified with FindVariableFeatures (method = “vst”). Group comparisons used a Wilcoxon rank-sum test, Welch’s t-test, and DESeq2 pseudobulk analysis. All p-values were corrected using Benjamini–Hochberg FDR. Effect sizes (Cohen’s d, Cliff’s Δ) and corresponding 95% CIs were calculated for all pseudobulk comparisons. For gene-level models, we used a linear mixed-effects model (lme4/lmerTest) with sample ID as a random effect to account for within-sample cell-level correlation. All analyses were scripted in R 4.3 with Seurat v5, edgeR/DESeq2, and ggplot2 v3.

### Statistical analysis

Unless otherwise specified, data are presented as mean ± standard deviation (SD). Comparisons between two groups were performed using Welch’s t-test (for normally distributed data) or the Wilcoxon rank-sum test (for non-normal data). When more than two groups were compared, one-way ANOVA with Bonferroni post hoc correction or the non-parametric Kruskal–Wallis test was applied as appropriate. For ddPCR copy-number assays and ddPCR-based MRGPRX2 expression measurements (HαT: n=16; non-HαT: n=19), group differences were evaluated using Welch’s t-test with effect sizes (Cohen’s d) and 95% confidence intervals reported.

Detailed statistical approaches for spatial transcriptomics and CyTOF analyses—including multiple-testing correction, effect-size calculations, and model structures—are provided in their respective Methods subsections. Briefly, spatial transcriptomic comparisons used DESeq2 with Benjamini–Hochberg false discovery rate (FDR) correction, and pseudobulk analyses were conducted at the sample level using both Wilcoxon and Welch’s t-tests, with effect sizes reported as Cohen’s d and Cliff’s Δ. Linear mixed-effects models (lme4/lmerTest) were used to account for within-sample cell-level correlation. For CyTOF datasets, arcsinh-transformed marker intensities were compared using Welch’s or Wilcoxon tests, with BH-FDR correction applied when multiple markers were evaluated simultaneously, and effect sizes (Cohen’s d, 95% CI) were calculated.

All statistical tests were two-sided. Given the exploratory nature and limited sample size of several cohorts, p-values should be interpreted with caution, and effect sizes and confidence intervals are emphasized throughout. Analyses were performed in GraphPad Prism (GraphPad Software), R (v4.3), and Python, using Seurat v5, DESeq2, lme4, Cytobank, and associated statistical packages.

## 3 Results

Tryptase genotyping was performed on 854 DNA samples from the University of Pennsylvania (n=433) and Mayo Clinic (n=421) IBD Biobanks using digital droplet PCR. Overall, 4.2% (36/854) of samples were positive for HαT (Figure 1A). Clinical stratification by IBD subtype (UC vs CD) was available for 433 samples. Within this subset, individuals with ulcerative colitis showed approximately twice the frequency of HαT (13/196) compared with those with Crohn’s disease (8/237) in the University of Pennsylvania cohort (Figure 1B).

To determine whether the genetic enrichment of HαT in IBD corresponded to detectable differences in mast-cell–associated pathways at the tissue level, we next examined transcriptomic and protein expression signatures in a series of nested molecular cohorts derived from the larger genotyped population. Because archived tissue availability varied across participants, each downstream analytic modality—spatial transcriptomics and CyTOF—was performed on the subset of samples for which high-quality RNA or viable tissue was accessible. These molecular analyses included matched HαT and non-HαT samples from individuals with well-phenotyped IBD (ulcerative colitis or Crohn’s disease), enabling us to evaluate whether α-tryptase gene dosage was associated with altered mast cell abundance, MRGPRX2 expression, or activation state within the intestinal mucosa. A summary of cohorts, tissue types, analytic platforms, and TPSAB1 copy-number distribution is shown in Table 1 to clarify the relationship between the genotyping dataset and downstream molecular experiments.

Next, we determined the expression of MRGPRX2 in these tissues using spatial transcriptomic analysis as well as ddPCR from 8 samples obtained from the biobank cohort - 4 with HαT and 4 without HαT from matching tissues, all descending colon with severe IBD at presentation. As expected, single-cell analysis of spatial transcriptomics sequencing data demonstrated higher numbers of MCs in the HαT samples. However, *MRGPRX2* expression was notably higher in MCs of HαT samples. This finding suggests that increased MRGPRX2 gene expression on HαT samples is not simply a function of more MC presence, but represents an increase in transcription (Figure 2A-C). We observed that MRGPRX2 expression was not limited to MCs, (Figure 2D), with relevant implications in subsequent studies. In addition, we observed that MRGPRX2 transcripts (red dots) are increased in the HαT tissues with IBD relative to the IBD alone group (Figure 2E). To validate the spatial transcriptomic findings, we analyzed RNA from a separate cohort of IBD with and without HαT by ddPCR and found that *MRGPRX2* gene expression was again significantly increased in HαT (Figure 2F). Pseudobulk analysis of the spatially sequenced data demonstrated that MRGPRX2 expression was globally increased in the HαT positive group, regardless of specific cell source (Figure 2G).

**Figure 2.**
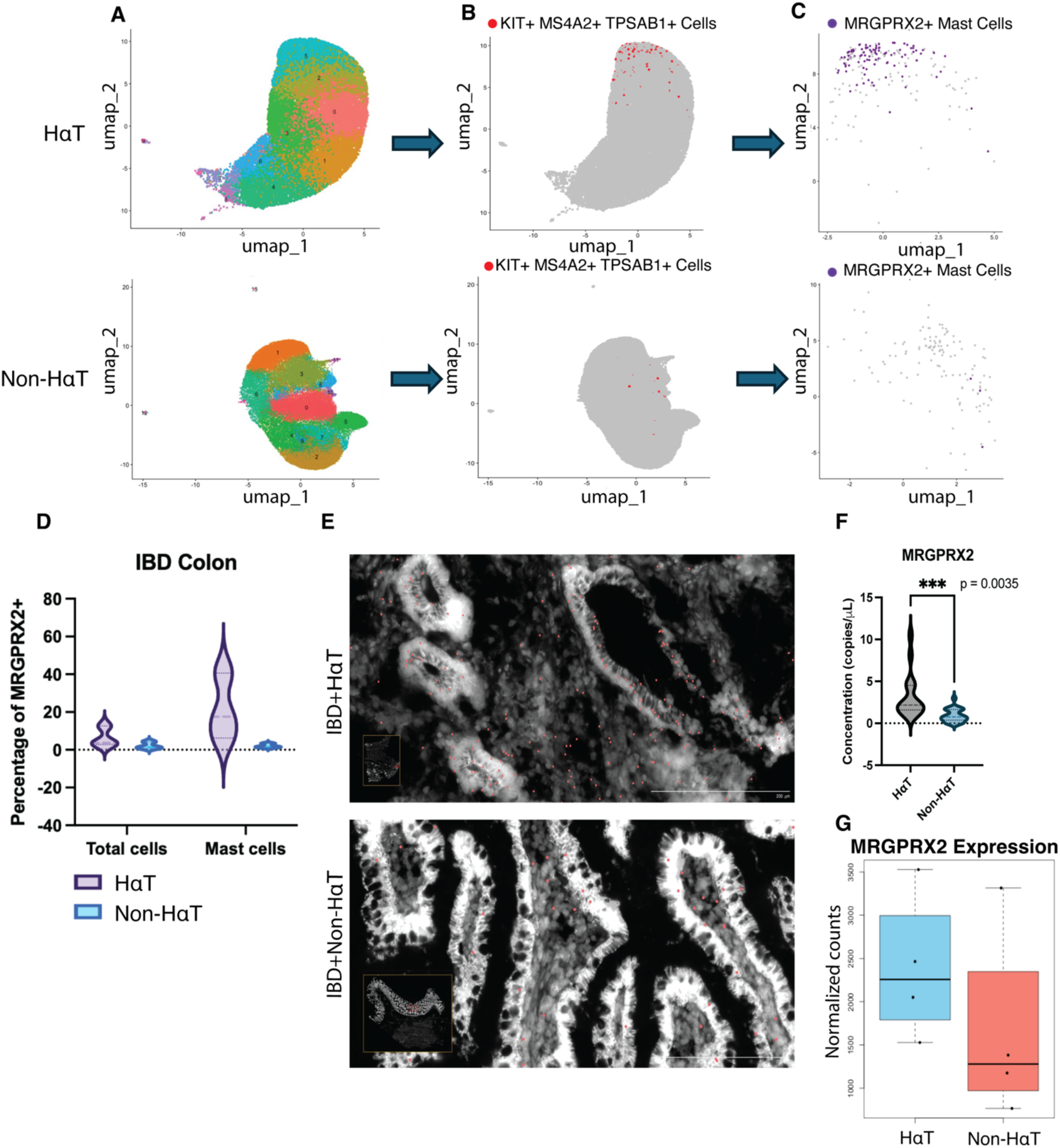
Mast cells from IBD patients with HαT demonstrate increased MRGPRX2 expression. Spatial transcriptomics (10x Xenium) was performed on 8 descending colon biopsies from the University of Pennsylvania IBD biobank (4 HαT, 4 non-HαT; balanced UC/CD). A) UMAP embedding showing major cellular populations. B) Mast cells (MCs), defined as TPSAB1⁺ MS4A2⁺ KIT⁺, are more abundant in HαT samples. C) Feature map of isolated MCs demonstrating increased MRGPRX2 transcript levels in HαT. D) Digital droplet PCR (ddPCR) of representative tissues from the same cohort confirms upregulated MRGPRX2 expression in HαT versus non-HαT. E) Spatial transcriptomics images showing increased MRGPRX2 transcripts (red dots) in HαT-positive IBD tissue compared with non-HαT tissue. F) ddPCR validation on matched samples (HαT: n=4; non-HαT: n=4) showing elevated MRGPRX2 mRNA. G) Pseudobulk differential expression demonstrates significantly increased MRGPRX2 in HαT samples. For transcriptomic analyses, differential expression was calculated using DESeq2 with Benjamini–Hochberg FDR correction (FDR < 0.05). Effect sizes are shown as log₂ fold-change with 95% CIs. For ddPCR comparisons, Welch’s t-test was used with Cohen’s d reported.

Next, to determine whether our observations in the colon were generalizable to other regions of the gastrointestinal tract, we analyzed mast cell (MC) phenotypes in the lamina propria of the small intestine using mass cytometry (CyTOF; antibody panel in Supplemental Table 2). The number of samples available for CyTOF analysis was inherently limited by the rarity of HαT and the challenge of obtaining duodenal tissue from symptomatic but histologically normal individuals undergoing endoscopy. Consequently, this component of the study represents a small exploratory cohort consisting of five HαT samples and twelve controls. These data are therefore intended to provide preliminary, hypothesis-generating insight into MC phenotypes rather than definitive estimates of effect size.

All small intestinal biopsies were histologically normal by hematoxylin and eosin staining. Two MC subpopulations (CD45⁺, Lin⁻, c-KIT⁺, FcεRI⁺, Tryptase⁺; gating strategy in Supplemental Figure 1) were identified. Across both non-HαT (Figure 3A) and HαT (Figure 3B) groups, MRGPRX2⁺ MCs demonstrated increased expression of activation-associated markers CD63, CD203c (ENPP3), LAMP1, and SIGLEC8 compared with MRGPRX2⁻ MCs. Notably, when examining the overall MC compartment independent of MRGPRX2 status, we did not observe significant differences in CD63, CD203c, or LAMP1 expression between HαT and non-HαT participants, suggesting that these changes are specific to the MRGPRX2⁺ MC subset. In contrast, SIGLEC8 expression was significantly increased in the HαT group (Figure 3C). While these findings provide initial evidence of distinct mast cell phenotypes associated with HαT in the small intestine, they remain exploratory and require validation in larger cohorts.

**Figure 3.**
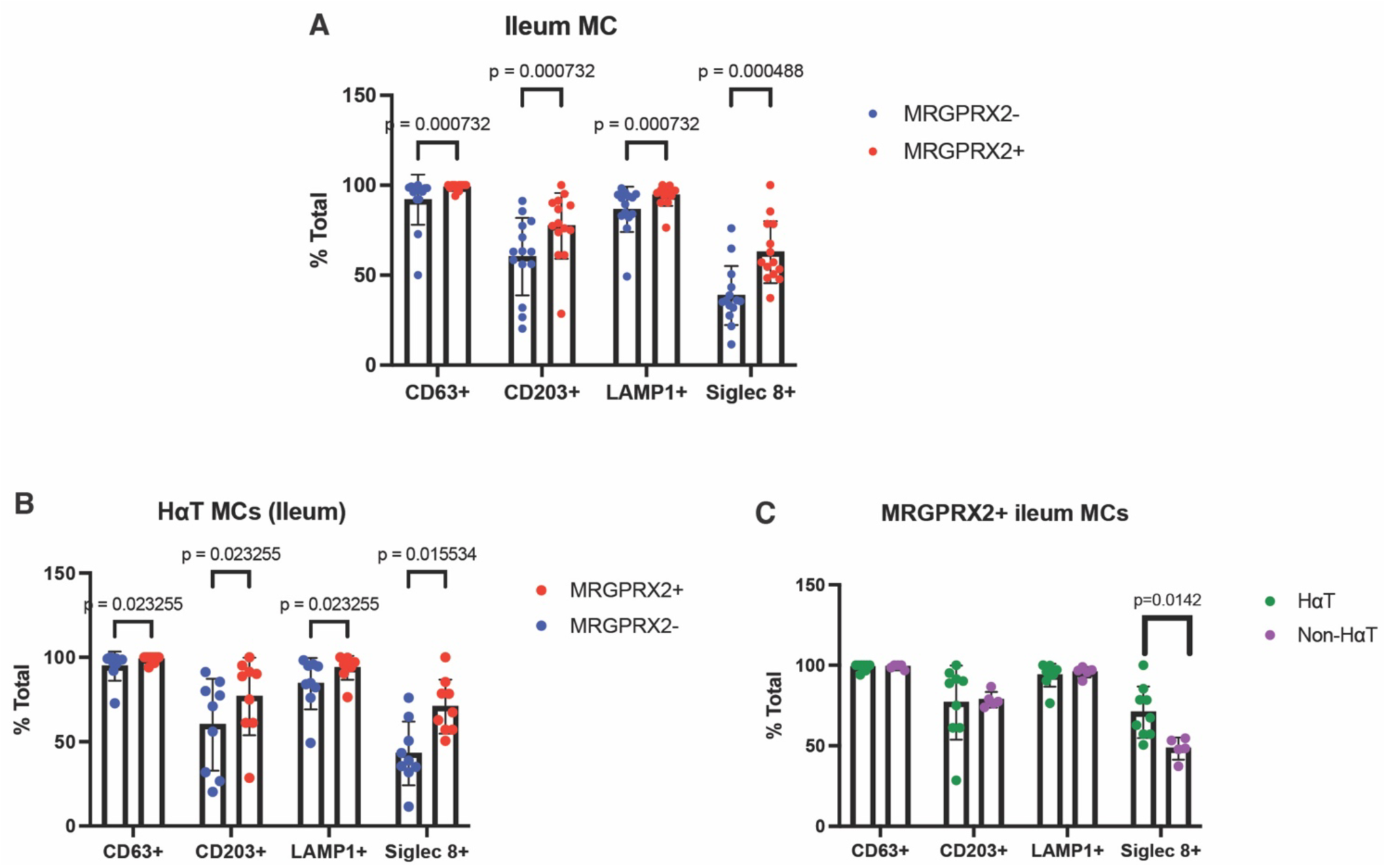
CyTOF analysis reveals increased SIGLEC8 in MRGPRX2⁺ mast cells within the small intestine of individuals with HαT. Small intestine biopsies from symptomatic, non-IBD participants (n = 17 total; HαT = 5, non-HαT = 9) were analyzed by mass cytometry (CyTOF). All samples were stained, barcoded, and acquired in a single run, with EQ bead normalization applied; no batch correction was required. Marker intensities were arcsinh-transformed (cofactor = 5). A) Across all samples, MRGPRX2⁺ MCs exhibited higher expression of CD63, CD203c, LAMP1, and SIGLEC8 compared with MRGPRX2⁻ MCs (Wilcoxon test; BH-FDR correction applied for multiple markers; effect sizes reported as Cohen’s d). B) In participants with HαT, MRGPRX2⁺ MCs showed significantly increased activation markers compared with MRGPRX2⁻ MCs. C) When comparing MRGPRX2⁺ MCs from HαT vs non-HαT individuals, SIGLEC8 was the only marker differentially expressed after FDR correction, indicating selective enhancement of SIGLEC8 in the HαT subgroup.

Finally, to further explore the expression of SIGLEC8, we performed pseudobulk analysis of the transcriptomic sequencing data obtained from the biobank IBD samples in order to determine differentially expressed genes per group (HαT vs non-HαT). Consistently, we found *SIGLEC8* was expressed at higher levels among patients with HαT (Figure 4A). When comparing HɑT vs non-HɑT samples by pseudobulk analysis, we also observed upregulation of genes related to epithelial integrity, such as *MS4A12*, *KRT1* and genes with immune regulatory functions (Figure 4B).

**Figure 4.**
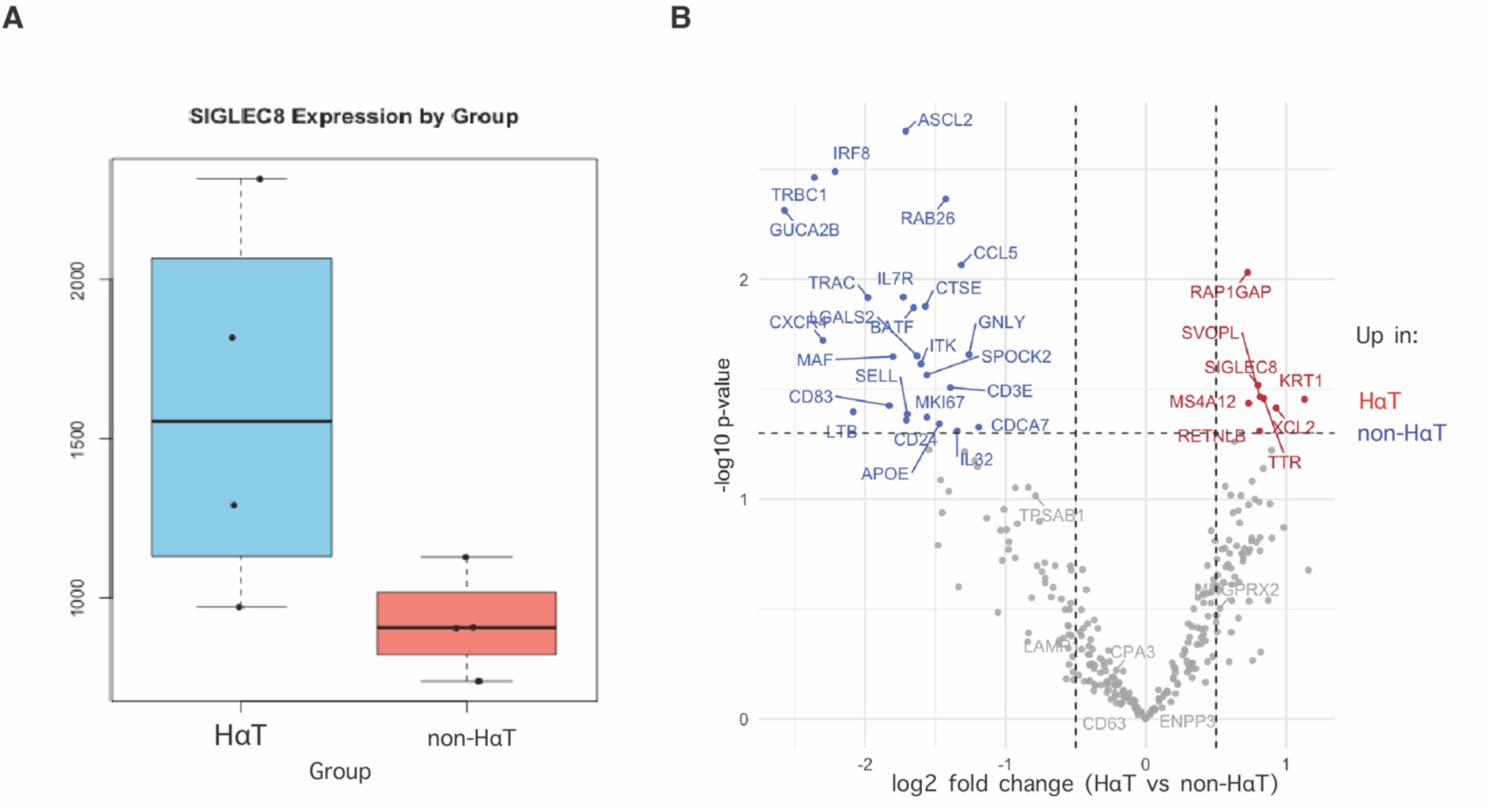
Individuals with IBD and HαT exhibit increased SIGLEC8 expression in colon tissue. Spatial transcriptomics and pseudobulk analysis were performed on 8 representative descending colon samples (4 HαT, 4 non-HαT; balanced UC/CD). A) Pseudobulk counts aggregated by sample show higher SIGLEC8 expression in the HαT group (Wilcoxon test; Cohen’s d and 95% CI reported). B. Volcano plot of DESeq2 pseudobulk differential expression analysis contrasting non-HαT (blue) and HαT (red) samples. Genes surpassing FDR < 0.05 (Benjamini–Hochberg correction) are highlighted. SIGLEC8 is prominently upregulated in HαT, consistent with findings from CyTOF and ddPCR validation.

## 4 Discussion

When UC and CD were considered together, the overall prevalence of HαT in our genotyped IBD population fell within the expected range for the general population (36). Although neither UC nor CD showed true enrichment, HαT occurred at approximately twice the frequency in UC compared with CD. Both values remain compatible with population variability, but the relative difference between UC and CD may warrant further exploration in larger cohorts. Within this framework, we observed increased MRGPRX2 expression in IBD tissue from individuals with HαT. Given prior links between MRGPRX2 and UC inflammation (25), and between HαT and gastrointestinal immunopathology (8), these data support the concept that HαT and MRGPRX2 expression may act as modifiers of IBD manifestations in at least a subset of patients.

Although mast cells are the principal source of MRGPRX2, we also observed MRGPRX2 transcripts outside mast cell clusters, suggesting that additional cell types within the intestinal mucosa may contribute to MRGPRX2-dependent signaling. The MRGPRX2⁺ mast cell subset showed increased expression of activation-associated markers, consistent with reports linking mast cell activation to IBD severity and activity (37,38). These findings are in keeping with an emerging view of mast cells as context-dependent regulators of mucosal inflammation rather than purely effector cells of allergy.

At the transcriptional level, non-HαT samples showed the expected high expression of T-cell–associated genes (TRBC1, TRAC, IL7R, CCL5, ITK, BATF), which have been linked to immunopathology in IBD (39–42). By comparison, HαT samples demonstrated comparatively reduced T-cell signatures and increased SIGLEC8 (43), consistent with a more mast-cell–skewed transcriptional program. Although prior clinical studies have reported a higher frequency of HαT among individuals evaluated for clonal mast-cell disorders, this association is likely influenced in part by referral patterns and baseline tryptase testing rather than a direct causal relationship. In this context, increased SIGLEC8 expression in our cohorts likely reflects counter-regulatory pathways and/or greater mast-cell abundance or activity (3,44,45), and may coincide with T-cell functional changes shaped by mast-cell mediators (46–48). Our exploratory findings suggest that HαT in IBD may be associated with heightened mast cell activity and upregulation of MRGPRX2 and SIGLEC8, pointing to a mast cell–centric endotype that could hold diagnostic or therapeutic relevance in refractory IBD. Although preliminary, these results modestly refine our understanding of mast cell states in the setting of elevated basal serum tryptase and underscore the potential clinical implications of mast cell–directed approaches in MC-driven disease (49,50)

Hereditary α-tryptasemia (HαT) has increasingly been recognized as a genetic modifier in mast cell (MC)–mediated conditions and is overrepresented in clonal MC disorders such as systemic mastocytosis (5,13,15,51). Foundational genetic studies demonstrated a gene-dose effect on basal serum tryptase (BST) together with an increased burden of mediator-related symptoms (3,15), supporting the concept that HαT can amplify MC-driven phenotypes in susceptible contexts. However, several groups have emphasized that the clinical impact of HαT is variable and may not define a single, uniform phenotype. For example, Chollet et al reported substantial heterogeneity among carriers in a small referral cohort (52), and Rama et al. (53) highlighted the broader diagnostic uncertainty that exists in mast cell activation disorders by demonstrating that commonly used clinical scoring systems have variable performance in predicting clonal disease. These findings underscore the complexity of interpreting mediator-related symptoms and BST elevations in clinical practice, and they reinforce the need to consider genetic contributors such as HαT alongside other sources of diagnostic ambiguity. Even so, two independent studies of idiopathic anaphylaxis (IA) without clonal mast cell disease found HαT in ∼17–29% of cases—far exceeding population rates—with increased TPSAB1 copy number accounting for persistently elevated BST in a subset of patients (12,54). Taken together, available evidence demonstrates that while HαT does not produce a uniform clinical syndrome, rather functions as a common genetic trait that increases susceptibility to severe or amplified MC-mediated responses under specific triggers or disease contexts. Supporting this model, duodenal biopsies from individuals with HαT show increased mucosal MC numbers compared with controls (8,16,33), indicating that α-tryptase gene dosage may influence tissue-level MC homeostasis and potentially modify disease expression beyond clonal mast cell disorders. Accordingly, the diagnostic uncertainty emphasized by these contrasting publications reinforce the importance of disentangling genetic factors—such as HαT—from other sources of variation in MC activation, a goal directly addressed by our study.

This study has some important limitations. First, the CyTOF component reflects a small exploratory cohort, driven by the practical challenges of obtaining duodenal biopsies from symptomatic but histologically normal individuals with HαT—a relatively uncommon genetic trait seldom biopsied outside research protocols. As a result, these findings should be interpreted as preliminary rather than definitive. Second, although we observed consistent trends in MRGPRX2 expression and mast-cell activation markers across datasets, larger cohorts will be required to determine the reproducibility and clinical significance of these associations.

Despite these constraints, our integration of genotyping, spatial transcriptomics, and high-dimentional immune phenotyping across independent cohorts reveals a reproducible MC-associated signature in individuals with HαT and IBD. These findings establish a biological foundation for future mechanistic studies and prospective clinical investigation. Our work highlights HαT as a potentially meaningful modifier of mucosal immunity in the GI tract.

*The authors declare that the research was conducted in the absence of any commercial or financial relationships that could be construed as a potential conflict of interest*.

## 5 Author Contributions

MGP performed ddPCR experiments, bioinformatics analysis and manuscript preparation. LK and DL performed CyTOF and CyTOF analysis. TR, ND, AC, and AO contributed by patient recruitment, processing patient samples, abstraction of patient data from the electronic health record. KW and GY performed ddPCR analysis and manuscript preparation. SS was involved in manuscript preparation. JJL established the index cohort, developed and performed genetic testing for HαT, contributed data related to supplemental Table 1, and was involved in data analysis and manuscript preparation. SCG was involved in patient recruitment, experimental design, data analysis, and manuscript preparation. HA was involved in experimental design, data analysis, and manuscript preparation.

## Data Availability

All data produced are available online at GitHub

https://github.com/nuzla/-GEO_HaT_Xenium_Submission

## Acknowledgments

The authors would like to acknowledge the University of Pennsylvania and Mayo Clinic IBD Biobanks for providing samples used in this study.

## Data Availability Statement

All code and processed data (expression matrices, CyTOF files, and metadata) are available in our public GitHub repository: https://github.com/nuzla/-GEO_HaT_Xenium_Submission. Additional materials may be requested from the corresponding author.

## Supplemental Figures and Tables

**Supplemental Table 1: Non-biobank cohort characteristics**

**Supplemental Table 2: Mass cytometry panel, clones, and sources.**

**Supplemental Table 3: R packages used for analysis**

**Supplemental Figure 1: Mass cytometry mast cell gating strategy.**

