## Supplementary material for "MRGPRX2-expressing mast cells are increased in the GI tract of individuals with active inflammatory bowel disease and hereditary α-tryptasemia": Sup tables

**Supplemental Table 1a.** Demographics of HαT Participants (N=8)

| Sample number | Sex at birth/race/ ethnicity | Diagnosis and TPSAB1 CNV | MC per HPF in small intestine | symptom onset | KIT | Serum tryptase | History of Anaphylaxis (Y/N) | Analysis |
|---|---|---|---|---|---|---|---|---|
| 5 | F/white/Caucasian | HAT 2:3 | 80 | Adult | Negative | 14.5 | Yes | C |
| 6 | M/white/Caucasian | HAT 3:2 | 32 | Adult | Negative | 17 | No | C |
| 7 | F/white/Caucasian | HAT 3:2 | 35 | Adult | Negative | 15.5 | Yes | C |
| 17 | M/white/Caucasian | HAT 2:3 | 75 | Adult | Negative | 12.8 | No | C |
| 18 | F/white/Caucasian | HAT 3:2 | 48 | Adult | Negative | 17 | Yes | C |
| 19 | M/white/Caucasian | HAT 2:3 | 35 | Pediatric | Negative | 10.7 | No | C |
| 20 | F/white/Caucasian | HAT 3:2 | 62 | Adult | Negative | 11.6 | yes | C |
| 21 | F/white/Caucasian | HAT 3:2 | 34 | Pediatric | negative | 10.6 | yes | C |

**Table 1b.** Demographics of Control Participants (N=4)

| Sample number | Sex at birth/race/ethnicity | TPSAB1 CNV | Serum Tryptase | analysis |
|---|---|---|---|---|
| 22 | F/white/Caucasian | 1:3 | N/A | C |
| 23 | F/white/Caucasian | | 5 | C |
| 24 | F/white/Caucasian | 0:4 | 4.1 | C |
| 25 | F/white/Caucasian | 0:4 | 4.7 | C |

**Supplemental table 2.** Mass cytometry panel, clones, and sources.

| Metal | General Panel | Clone | Place of Purchase |
|---|---|---|---|
| 89 | CD45 | HI30 | Fluidigm |
| 115 | CD34 | 581 | Core, self conj |
| 141 | c-kit | *104D2* | core, self conj |
| 142 | CD19 | HIB19 | Core |
| 143 | HLA-DR | L243 | Fluidigm |
| 144 | CD64 | 10.1 | Core |
| 145 | cRTh2 | BMI6 | core, self conj |
| 146 | CD8a | RPA-T8 | Core |
| 147 | CD45RO | UCHL1 | Core |
| 148 | CD28 | CD28.2 | Core |
| 149 | CD25 | 2A3 | Fluidigm |
| **150** | **CD63** | **H5C6** | **Fluidigm** |
| 151 | CD2 | TS1/8 | Fluidigm |
| 152 | CD14 | M5E2 | Core |
| 153 | CD45RA | HI100 | Core |
| 154 | CD33 | WM53 | Core |
| 155 | CD27 | L128 | Fluidigm |
| 158 | CD3 | UCHT1 | Core |
| 159 | CD11c | Bu15 | Core |
| **160** | **MRGPX2** | 47753 | **core conjugated** |
| 161 | CD203c | NP4D6 | core, self conj |
| 162 | CD16 | 3G8 | core, self conj |
| 163 | CD183(CXCR3) | G025H7 | Fluidigm |
| **164** | **Siglec 8** | **7C9** | **Fluidigm** |
| 165 | Tryptase | AA1 | core custom |
| **166** | **Lamp-1** | **H4A3** | **Core** |
| 168 | CCR6 | G034E3 | Core |
| **169** | **CD24** | **ML5** | **Fluidigm** |
| 170 | CCR7 | G043H7 | Core |
| 171 | CD127 | A019D5 | Core |
| 172 | IgM | MHM-88 | Core |
| **173** | **FCeR1** | **AER-37** | **Core** |
| 174 | CD4 | SK3 | Fluidigm |
| 175 | CD66b | G10F5 | Core, custom conj |
| 176 | CD56 | *N901* | *fluidigm* |
| 209 | CXCR5 | MU5UBEE | Core, custom conj |

**Supplemental table 3.** R Packages used in transcriptomics analysis

| Package | Version | Source |
|---|---|---|
| **R** | 2023.06.1+524 | CRAN |
| **Seurat** | v5.3.0 | CRAN |
| **SCTransform (in Seurat)** | 0.4.2 | CRAN |
| **ggplot2** | 3.5.2 | CRAN |
| **cowplot** | 1.1.3 | CRAN |
| **Matrix** | 1.6-4 | CRAN |
| **stringr** | 1.5.1 | CRAN |
| **DESeq2** | 1.42.1 | Bioconductor |

**Supplemental Figure 1.** CyTOF gating strategy

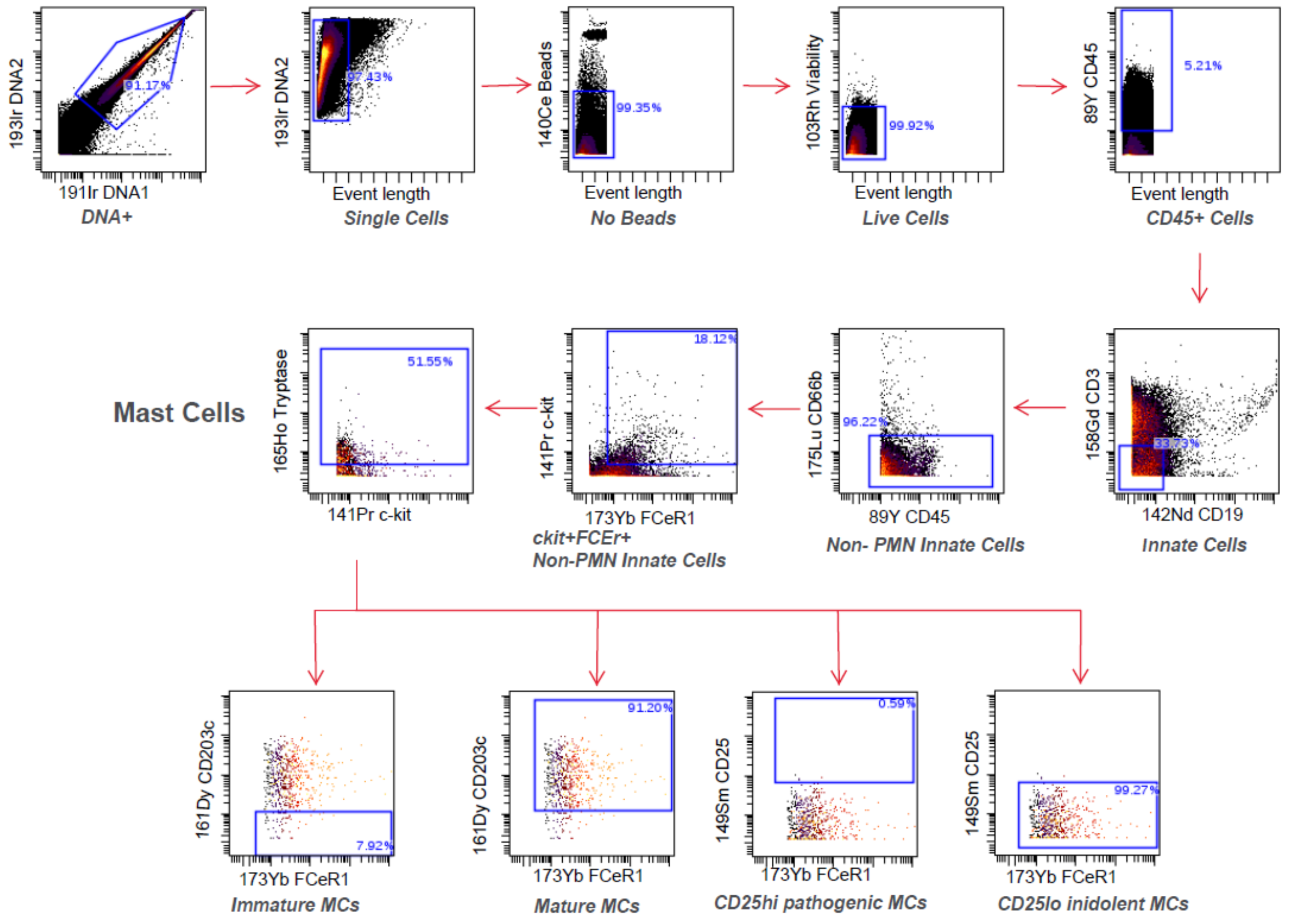
